# Comprehensive Analysis of the Human Urinary Peptidome Using Peptide Clustering Approach Reveals Age- and Gender-Specific Patterns

**DOI:** 10.1101/2025.05.16.25327757

**Authors:** Amr Elguoshy, Keiko Yamamoto, Yoshitoshi Hirao, Tomohiro Uchimoto, Kengo Yanagita, Tadashi Yamamoto

## Abstract

**Background:** The human urine peptidome reflects physiological and pathological states, making it a valuable resource for biomarker discovery. However, endogenous peptides often exist as cascades of truncated variants, complicating comparative analyses. To address this, we developed a “peptide cluster” approach, grouping overlapping peptides into representative clusters for robust statistical evaluation.

**Methods:** Urine samples from 55 healthy volunteers (23 males, 32 females) were analyzed via LC-MS/MS. Identified peptides were assembled into clusters based on sequence overlap, with the longest peptide designated as the “precursor” and truncated variants as “truncated“

**Results:** We identified 30,471 endogenous peptides, assembled into 13,163 peptide clusters—the largest urinary peptidome dataset to date. Gender-specific differences were observed in 26 clusters, while 57 clusters correlated significantly with age. Notably, male-enriched clusters included hepcidin-25 and progranulin-derived peptides, whereas female-enriched clusters were linked to immunoglobulin gamma-1. Age-associated clusters highlighted collagen degradation patterns, consistent with extracellular matrix remodeling.

**Conclusion:** Our peptide clustering approach facilitated a comprehensive characterization of the endogenous peptidome, capturing the diversity of naturally occurring truncated peptide forms. The resulting age- and sex-specific peptide clusters serve as a valuable reference framework for future investigations into disease-associated biomarkers.

## Introduction

Endogenous peptides, the naturally occurring endogenous peptides present in biological samples, are now comprehensively identified and characterized through peptidomics approaches [1–3]. These peptides, typically of low molecular weight, are generated by proteolytic cleavage of precursor proteins, often in conjunction with post-translational modifications (PTMs) [2]. Endogenous peptides play diverse and critical biological roles, functioning as hormones, cytokines, neuropeptides, and growth factors. Their generation is tightly regulated by proteolytic systems that maintain cellular homeostasis, ensuring the balance between protein synthesis and degradation [2,3].

Intracellular protein turnover is primarily governed by two key pathways: the ubiquitin-proteasome system (UPS)—the principal mechanism for selective protein degradation [4]—and autophagy, where proteins are sequestered and delivered to lysosomes for enzymatic breakdown [5]. Within these systems, cysteine, aspartate, and threonine proteases function as the main intracellular endopeptidases at acidic pH. In contrast, extracellular protein degradation is mediated by metalloproteinases and serine proteases, which operate at neutral pH within the extracellular matrix[6] [7]. Disruption of these finely tuned proteolytic processes can contribute to disease onset and progression. The truncated peptide forms generated by proteases thus reflect the dynamic proteolytic landscape of the organism and serve as molecular signatures of physiological and pathological states.

Among biological fluids, urine offers a particularly valuable window into the body’s peptidomic profile. Urinary peptides are predominantly derived from the kidney and urinary tract, providing detailed insights into diseases affecting these systems. A smaller fraction originates from the systemic circulation, offering additional clues about systemic physiological states [8]. Importantly, urine collection is non-invasive, and unlike blood, the urinary peptidome is relatively stable. Proteolytic processes are believed to be largely complete by the time urine is voided, meaning that urinary peptides represent end-products of proteolysis, providing a snapshot of the in vivo peptidomic state without further degradation [9].

Comprehensive profiling of the urine peptidome in healthy individuals is invaluable, not only for understanding the normal physiology of the kidney and urinary tract but also for capturing systemic homeostasis. Deviations from the baseline peptidomic profile can signal disease processes in both renal and non-renal organs.

Although extensive research has explored proteomic profiles across various human tissues and fluids, relatively few studies have systematically examined the human peptidome, which holds great potential for biomarker discovery and for uncovering bioactive peptides and proteolytic mechanisms. Early peptidomic studies have been largely limited to specific cohorts or conditions. For instance, Ashley et al. identified 4,543 endogenous urinary peptides from 566 precursor proteins in healthy individuals using liquid chromatography– mass spectrometry (LC-MS) [8]. Similarly, Julie et al. analyzed the urinary peptidome in youths with type 1 diabetes and controls, identifying 6,550 peptides derived from 751 protein precursors and highlighting a signature of uromodulin peptides linked to early diabetic changes[10].

However, in these studies, endogenous peptides were analyzed using workflows originally developed for tryptic peptides in proteomics. This approach overlooks a key feature of endogenous peptides: their natural variability due to protease activity, often differing only by the addition or removal of a few amino acids at the N- or C-terminus. Such peptides form cascades, where shorter truncated peptide forms are derived from longer peptides by amino- or carboxypeptidases.

To address this, we developed a novel peptidomics approach based on peptide clustering. In this method, overlapping peptides are assembled into unified structures called “peptide clusters,” each comprising a longest “precursor peptide” and its truncated derivatives, termed “truncated peptides.” This clustering strategy captures the dynamic proteolytic processing of peptides more accurately than conventional analysis.

Despite the promise of urinary peptidomics, few studies have examined the influence of fundamental biological variables—such as age and gender—on the urinary peptidome under normal physiological conditions, and none have done so using a peptide cluster-based approach.

In this study, we aim to systematically characterize the human urinary peptidome under normal physiological conditions using our peptide clustering methodology. Furthermore, we investigate age- and gender-related variations in the urinary peptidome, providing a robust reference framework for future biomarker discovery efforts that consider demographic variables in health and disease contexts.

## Results and Discussion

### Comprehensive Profiling of the Urine Peptidome

In total, we identified 30,471 endogenous peptides derived from 7,669 unique precursor protein groups across 55 urine samples (23 males, 32 females), using the Mascot search engine at a stringent 5% PSM FDR. To the best of our knowledge, this represents the most extensive peptidome coverage reported to date, significantly expanding the current landscape of urinary peptide biomarkers.

Given the intrinsic property of endogenous peptides to differ by minor truncations at the N- or C-termini, we systematically grouped overlapping peptides into distinct clusters based on sequence alignment. In this framework, the longest, complete sequence within each cluster is designated the “precursor peptide,” while shorter, overlapping sequences are termed “truncated peptides” (Figure 1). Notably, these precursor peptides were either directly identified or inferred through reconstruction from their truncated peptide forms. Through this clustering approach, we consolidated the 30,471 peptides into 13,163 distinct peptide clusters.

**Figure 1.**
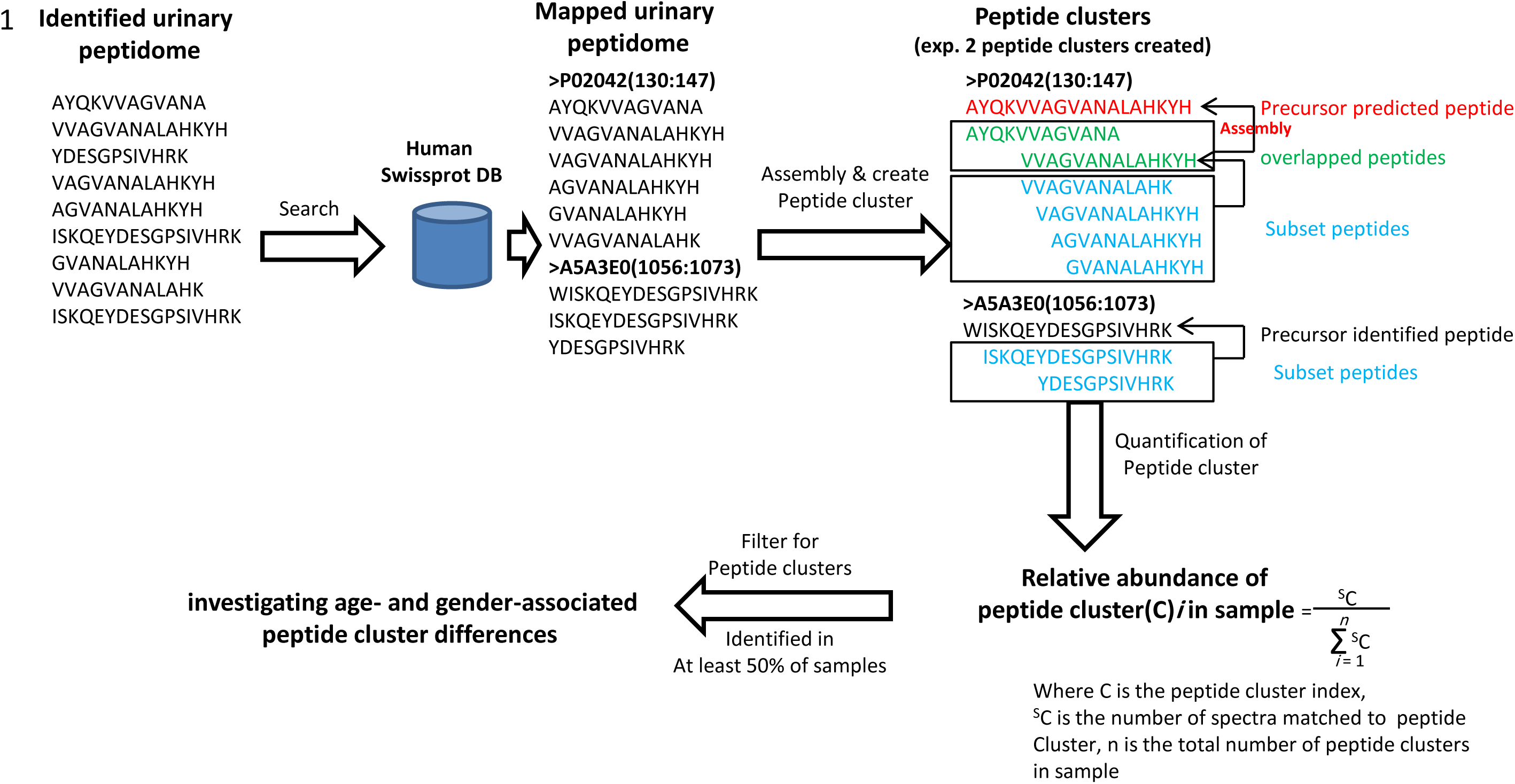
Peptidome analysis workflow illustrating the assembly of overlapped endogenous peptides and building the “peptide cluster”.

### Structural Organization of the Peptidome: Clustering Dynamics

Examining the distribution of peptide clusters revealed that approximately 35.5% of precursor proteins gave rise to multiple distinct clusters, with a clear inverse relationship between cluster frequency and cluster count per protein (Figure 2A). Importantly, protein length showed a statistically significant positive correlation with the number of peptide clusters generated (R = 0.33, *P* < 2.2e-16), suggesting that larger proteins serve as richer substrates for proteolytic diversification (Figure 2B).

**Figure 2.**
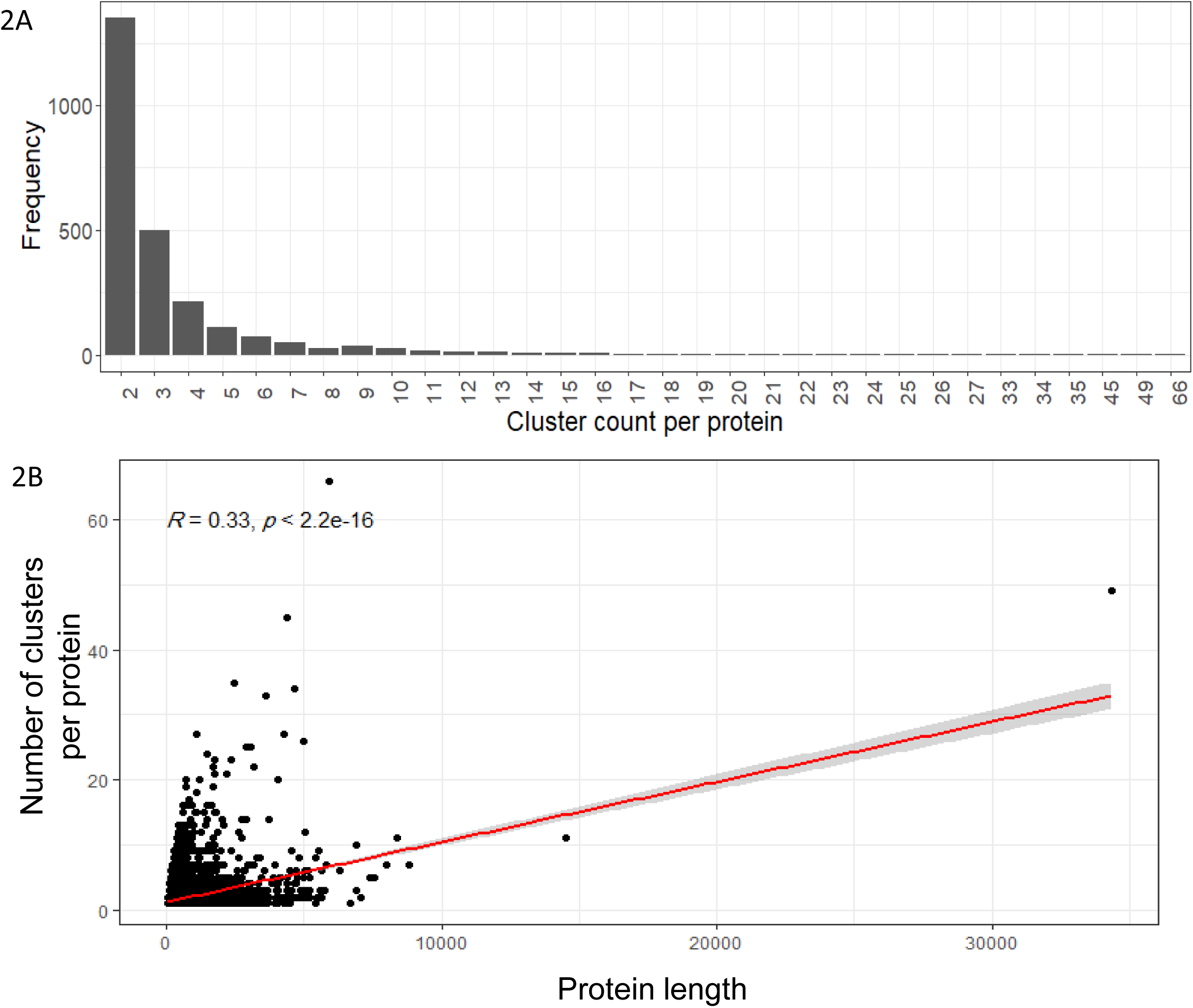
**A**: Bar plot showing the distribution of peptide cluster through precursor proteins., **B:** Correlation plot showing a significant positive correlation (R = 0.33, p < 2.2e-16) between protein length and peptide cluster count.

Furthermore, analysis of peptide fragmentation patterns within clusters revealed that about 33% of precursor peptides underwent further truncation into one or more truncated peptide forms. Longer precursor peptides were particularly prone to this process, as evidenced by a strong positive correlation between precursor length and fragment count (R = 0.73, *P* < 2.2e-16; Figure 3B). Moreover, clusters with more truncated peptide forms tended to be more abundant, as indicated by the significant positive correlation between the count of truncated peptide forms and cluster abundance (R = 0.67, *P* < 2.2e-16; Figure 3C).

**Figure 3.**
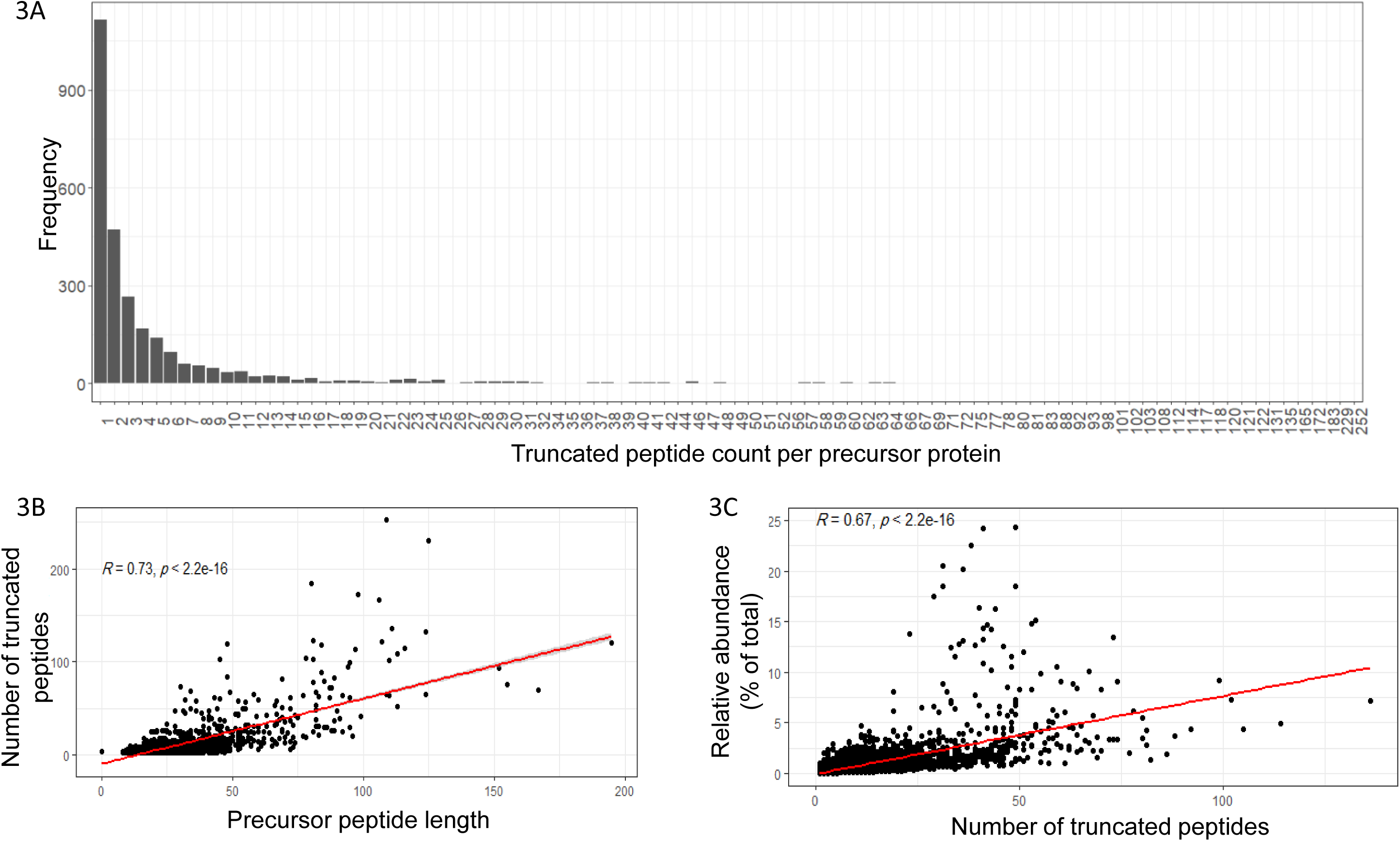
**A:** Bar plot showing the distribution of the truncated peptide across different precursor peptides., **B:** Correlation plot showing a significant positive correlation (R = 0.73, p < 2.2e-16) between precursor peptide length and truncated peptide count., **C:** Correlation plot showing a significant positive correlation (R = 0.67, p < 2.2e-16) between the number of truncated peptides and relative abundance as percent of total.

Remarkably, though precursor peptides accounted for only 4% of the total peptidome signal, their truncated peptide forms contributed a substantial 88%, highlighting the dominance of truncated peptides in the urinary peptidome. As illustrative cases, a fibrinogen alpha chain precursor spanning residues 289–413 yielded 229 distinct truncated peptide forms, while uromodulin precursor (residues 568–612) produced 102 truncated peptide forms.

Conversely, approximately 67% of precursor peptides remained un-truncated, representing 7.6% of the total signal. These intact precursors appear to cluster in specific protein regions, hinting at potential bioactivity or inherent stability that may confer resistance to proteolysis. Notably, 19 peptide clusters, such as the insulin C-peptide cluster, were detected universally across all samples, underscoring their potential as robust biomarkers.

### Sex-Specific Signatures in the Urinary Peptidome

We next examined gender-specific peptide cluster patterns. Common clusters were more consistently detected across samples, whereas sex-specific clusters appeared in fewer individuals (Figure 4). For instance, cluster 166, originating from the protease inhibitor SERPINB3 (residues 305–382), was identified exclusively in 16 female samples, aligning with prior findings that SERPINB3 is enriched in female-specific tissues and urine [11]. Intriguingly, 9 out of 11 clusters from SERPINB3 displayed this female specificity.

**Figure 4.**
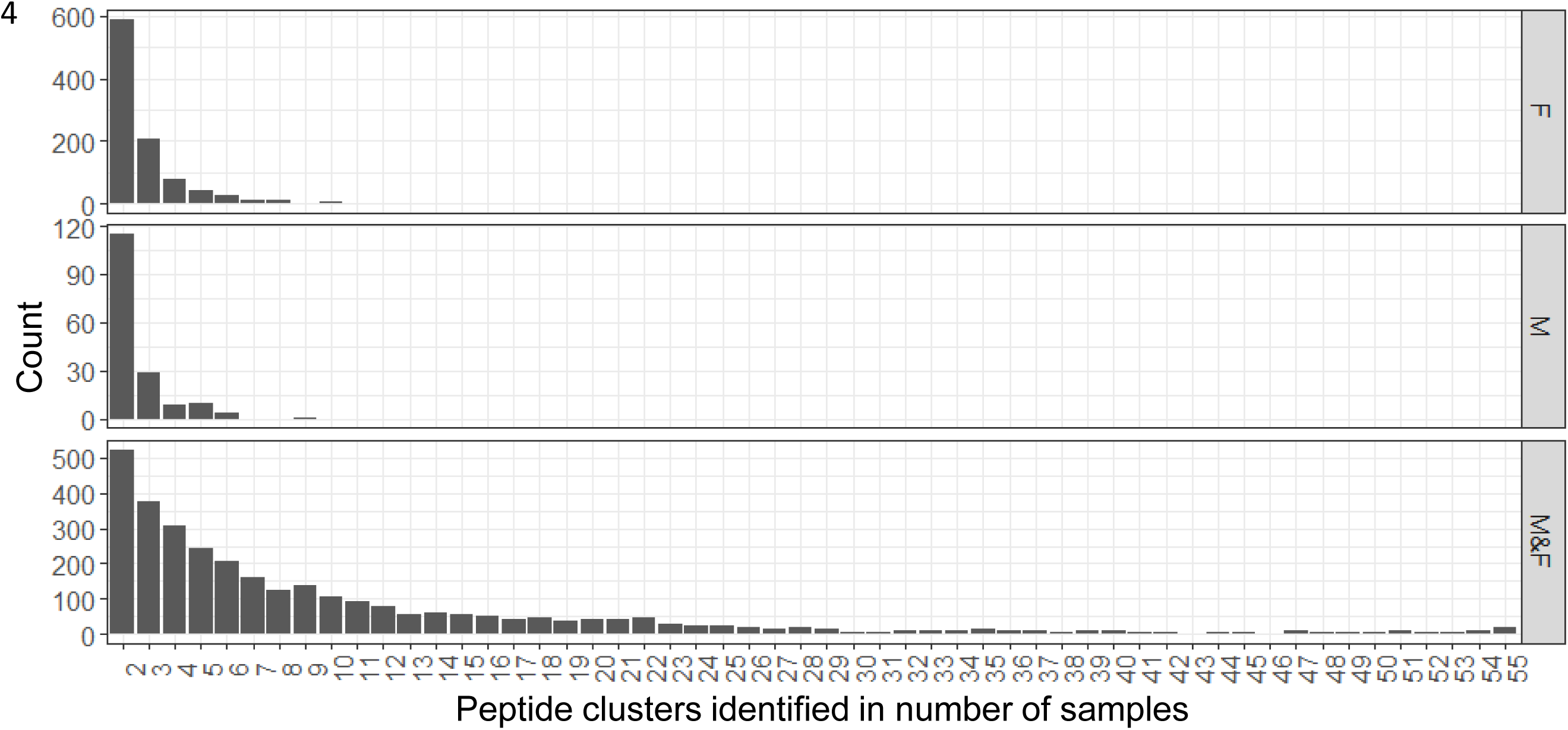
Bar plot showing the peptide clusters identified in number of samples

In contrast, male-specific clusters were exemplified by cluster 77, derived from prostate-specific antigen (PSA; residues 75–169), which appeared in 9 male samples. This cluster included one precursor and 20 distinct truncated peptide forms, with expression consistent with PSA’s tissue specificity according to the Human Protein Atlas.

To focus subsequent analyses on robust signals, we filtered for clusters detected in at least 50% of samples, yielding 199 peptide clusters for differential quantification.

### Stability Dynamics of Precursor Peptides

Despite peptides’ inherent vulnerability to enzymatic degradation [12], emerging data suggest that some exhibit remarkable stability, attributed to intramolecular forces such as hydrophobic interactions, hydrogen bonds, and disulfide bridges that reduce entropy [13]. To assess this, we calculated a *stability index* for each precursor peptide in our core set (n = 199), integrating spectral counts and peptide length:

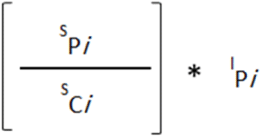

Where Pi is the Precursor peptide index and Ci is the peptide cluster index,

^S^P is the number of spectra matched to precursor peptide i

^S^C is the number of spectra matched to peptide cluster i

^l^Pi is the length of the precursor peptide Pi

Using the HLP web server[14], we predicted peptide half-life and entropy as stability indicators. Strikingly, peptides with higher SI values corresponded to longer predicted half-lives and lower entropy (Figure 5A–B), supporting the hypothesis that these peptides are inherently more resistant to urinary peptidases. This trait may explain their persistent detectability across samples and points to their potential as stable biomarkers.

**Figure 5.**
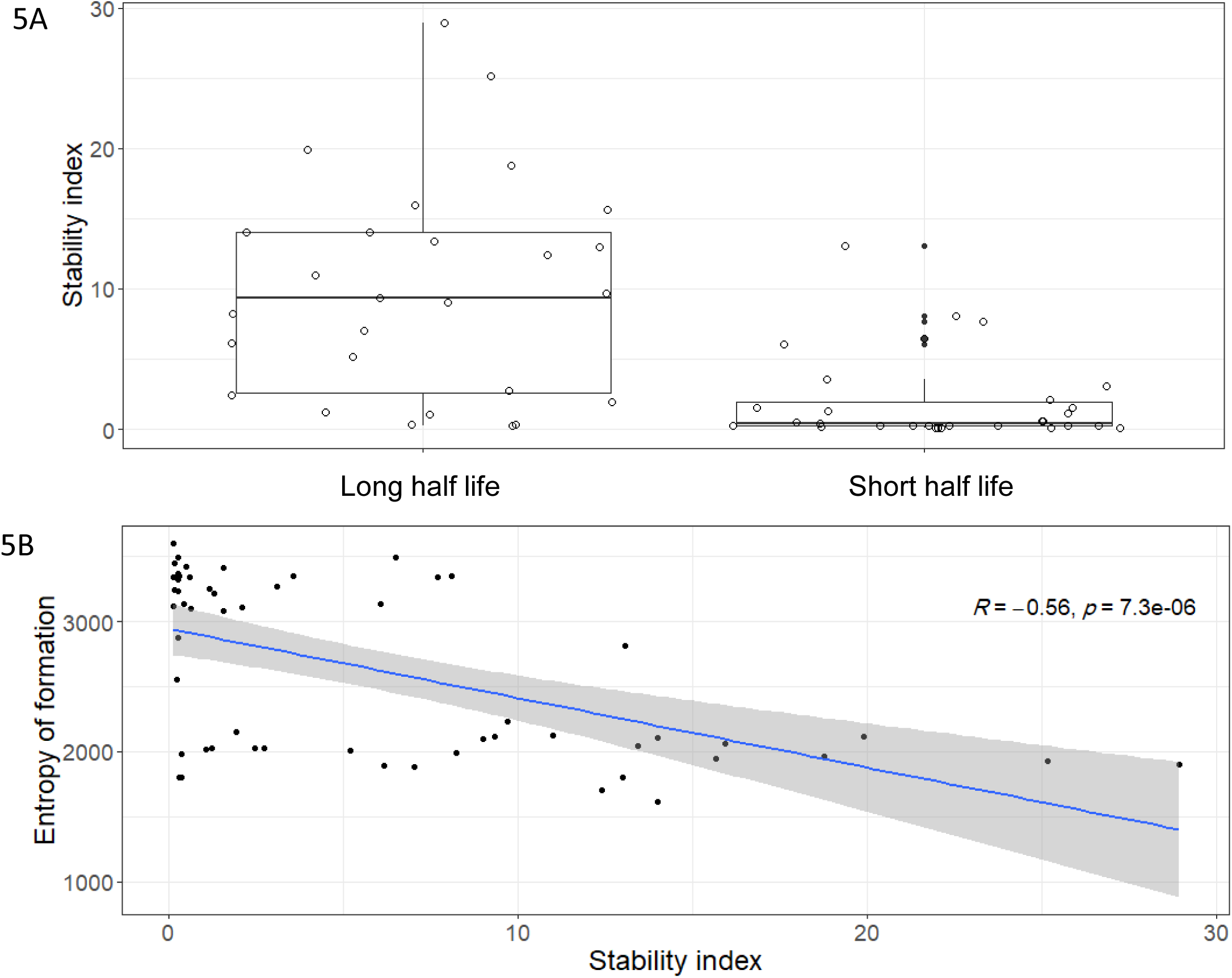
**A**: Boxplot showing the distribution of the stability index values in long and short half-life precursor peptide groups., **B.** scatterplot showing negative correlation between stability index values and Entropy.

### Investigation of the differentiated peptide clusters between male and female

Differential analysis between males and females revealed 26 out of the 199 clusters were significantly altered (≥1.5-fold change, *P* < 0.05), with 7 upregulated and 19 downregulated in females (Figures 6A, B, C) (Table S1). Importantly, several of these clusters correspond to biologically meaningful proteins with known sex-dependent expression.

**Figure 6.**
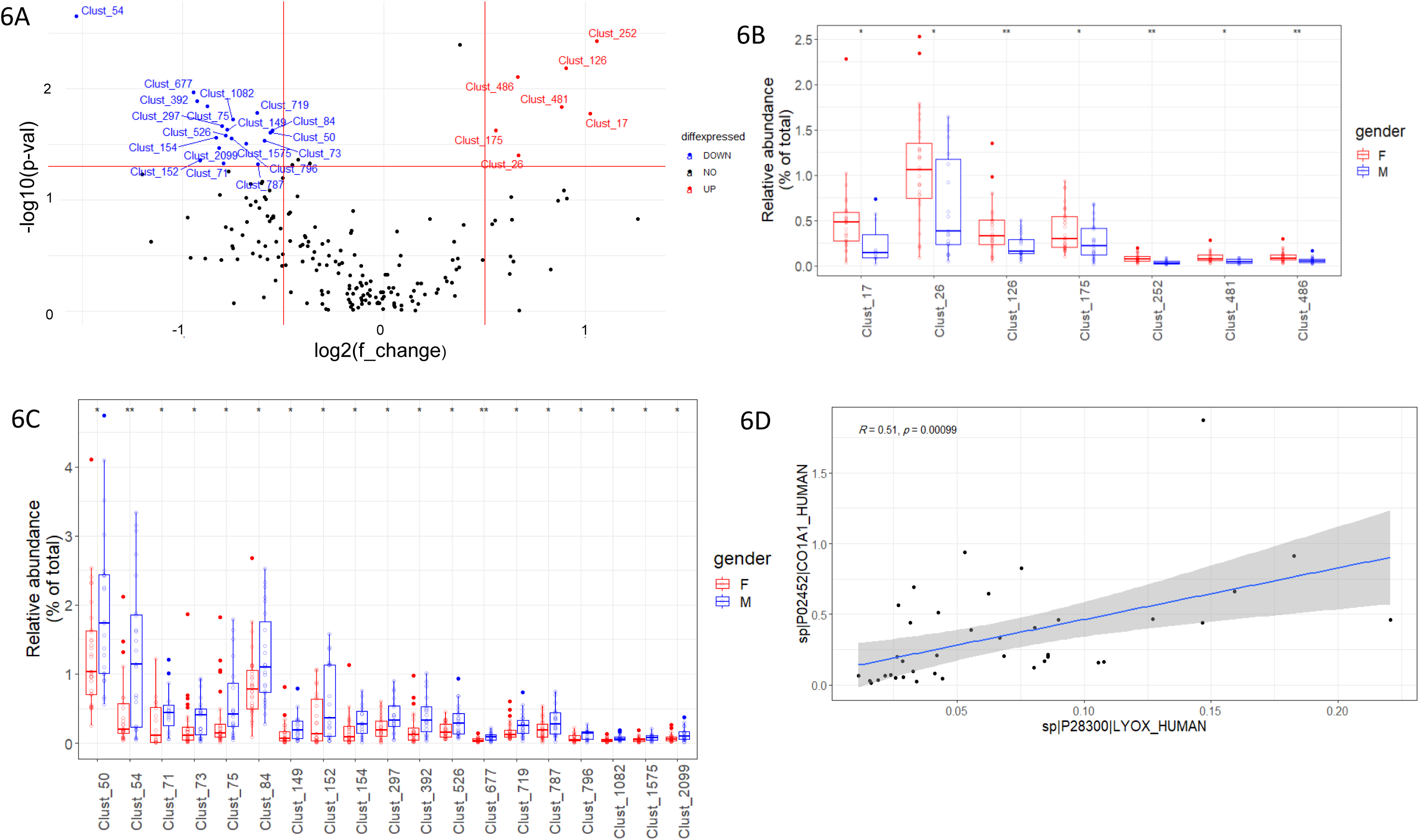
**A:** Volcano plot showing the differentiated peptide clusters between male and female., **B:** Bar plot showing the upregulated peptide clusters in females than males. **C:** Bar plot showing the upregulated peptide clusters in males than females. **D:** Correlation plot showing a significant positive correlation (R = 0.51, p < 0.00099) between the expression of clust_677 in Lysyl oxidase (LYOX) protein and clust_73 in collagen-alpha1 chain.

For example, cluster 54, the most significantly upregulated in males, corresponds to hepcidin-25—a bioactive 25-residue peptide “DTHFPICIFCCGCCHRSKCGMCCKT” that regulates iron homeostasis through ferroportin degradation [15]. Consistent with our findings, serum hepcidin levels have been reported to be significantly higher in males [16].

Similarly, two upregulated clusters in males originated from progranulin, a multifunctional secreted protein implicated in neurodegenerative diseases [17]. Cluster 50 (residues 17–47) and a second cluster (residues 279–337) were highly prevalent and truncated, supporting their biological relevance. Notably, increased progranulin expression in males may contribute to their lower susceptibility to Alzheimer‘s disease, as previously suggested [18].

Gene ontology analysis of the precursor proteins associated with male-upregulated peptide clusters revealed significant enrichment in the biological process of collagen fibril organization. Three proteins—collagen alpha-1(I) chain, collagen alpha-1(XVIII) chain, and lysyl oxidase (LYOX)—were prominently enriched in this category.

In the collagen alpha-1(I) chain, two peptide clusters (clust_73 and clust_719) were detected in 53 and 54 samples, respectively. Clust_73 corresponds to a 28-residue peptide (residues 1168–1195), generating 29 distinct truncated peptides. Clust_719 comprises a 10-residue peptide (residues 1197–1206), yielding 3 truncated peptides. Both peptides are located at the C-terminus of the protein. Similarly, clust_84 in the collagen alpha-1(XVIII) chain was identified in all 55 samples and corresponds to a 24-residue peptide (residues 1534–1557) truncated into 25 truncated peptide forms. Notably, both collagen alpha-1(I) and alpha-1(XVIII) are major constituents of the interstitial matrix, contributing to its structural integrity.

The lysyl oxidase (LYOX) protein, which catalyzes the formation of covalent cross-links in collagens and elastin[19], also exhibited a sex-biased peptide cluster. Peptide cluster 677, derived from the N-terminal propeptide region (residues 24–45) of LYOX, was detected in 39 of 55 samples and yielded 4 truncated peptides. According to UniProt, this region is part of the lysyl oxidase propeptide domain (residues 22–186).

Given the integral role of lysyl oxidase in collagen cross-linking, we tested for correlations between its peptide cluster and those from collagen proteins. A significant positive correlation was observed between LYOX cluster 677 and collagen alpha-1(I) cluster 73 (R = 0.51, p = 0.00099; Figure 6D), suggesting a co-regulatory relationship or functional interplay during extracellular matrix remodeling.

On the other hand, the 19 peptide clusters downregulated in males (thus upregulated in females) including Immunoglobulin gamma-1 heavy chain, albumin, actin, cytoplasmic 1, Thy-1 membrane glycoprotein, Golgi apparatus protein 1, multimerin-2, and podocalyxin (Figure 6A, B) (Table S1).

The precursor peptide in Immunoglobulin gamma-1 heavy chain was 97 aa peptide at the region between 115 and 211. It was truncated to 113 distinct peptides. It was reported in previous study conducted on mice that females generally showed higher levels of antibodies than males and it was statistically significant for IgG subclasses, explaining the reason behind increasing the severity of influenza virus disease in men compared to women[20].

### Investigation of age-correlated urinary peptide clusters

To investigate the potential relationship between ageing and the urinary peptide landscape, we assessed the correlation between the relative abundances of 199 peptide clusters and donor age. This analysis identified 57 peptide clusters significantly associated with age (p ≤ 0.05) (Tables S2–S4).

Strikingly, the majority of these ageing-associated peptide clusters exhibited sex-specific associations. Of the 57 significant clusters, 50 (88%) were associated with age in males, while only 4 clusters (7%) were associated with age in females. The remaining 3 clusters (5%) showed age correlations that were independent of sex (Supplementary Table S5).

Among the 50 age-associated peptide clusters in males, 38 displayed a positive correlation with age, suggesting an age-dependent increase in abundance. Conversely, 11 clusters were negatively correlated with age (Figure 7A) (Table S2). These findings suggest that ageing in males is characterized by a broader shift in the urinary peptidome compared to females.

**Figure 7.**
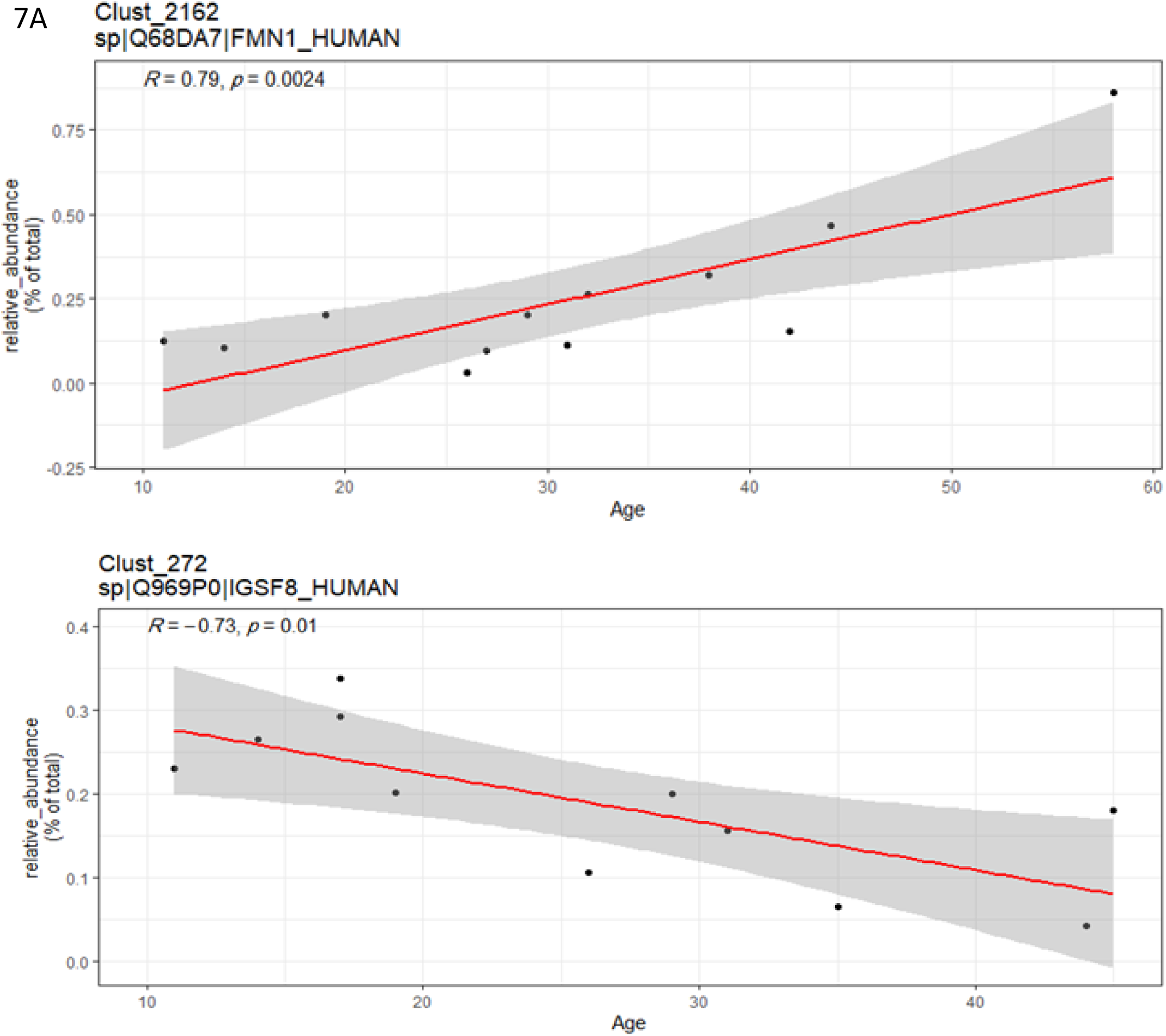
**A:** Correlation plot showing examples of age-correlated urinary peptide clusters with a significant positive or negative correlation (p < 0.05) in males.

In contrast to males, only four peptide clusters were significantly correlated with age in females. Of these, two showed positive and two showed negative correlations with age (Figure 7B) (Table S3). The limited number of ageing-associated peptides in females may reflect differences in biological aging processes or renal peptide processing and excretion between sexes.

**Figure 7.**
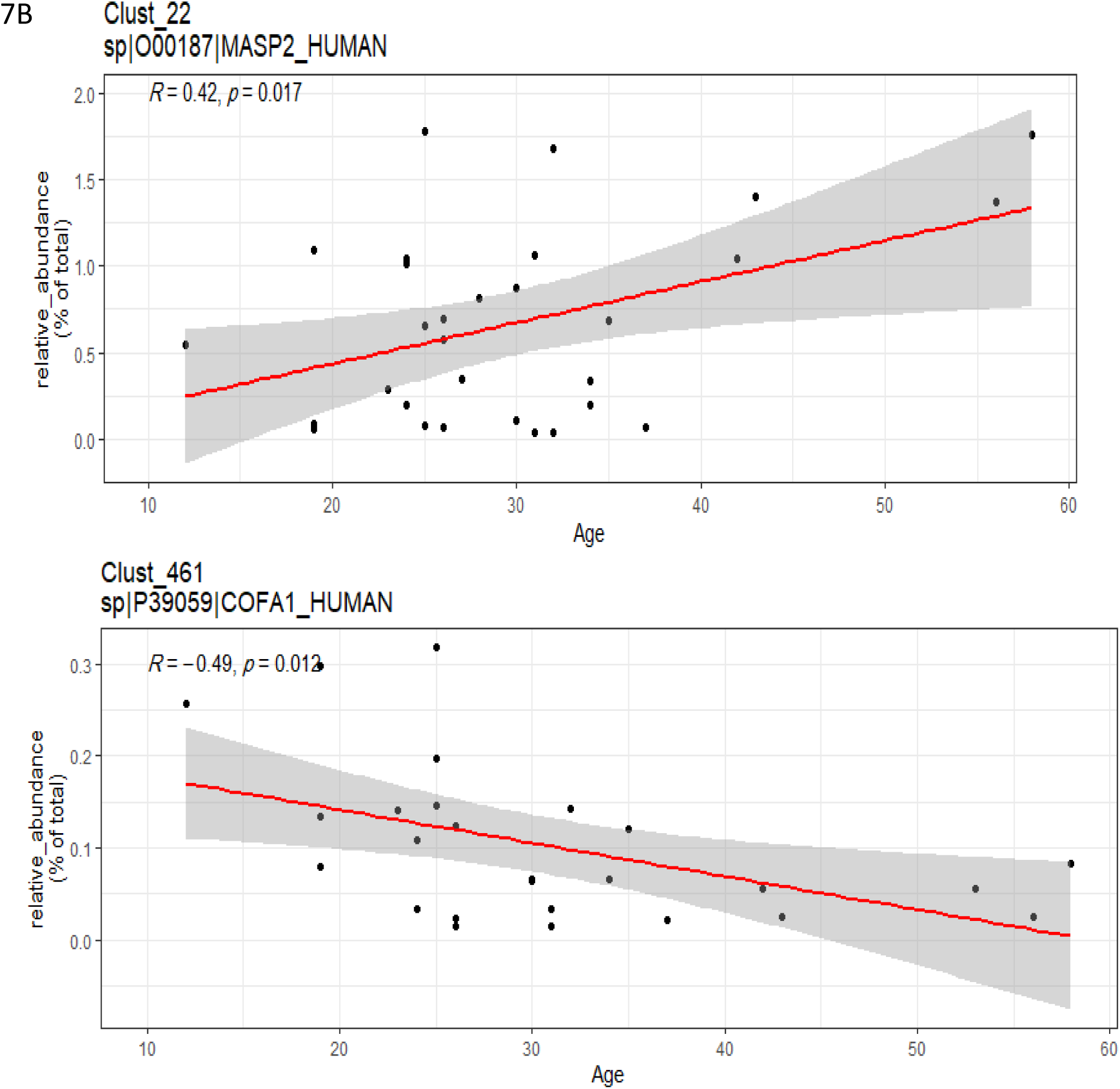
**B:** Correlation plot showing examples of age-correlated urinary peptide clusters with a significant positive or negative correlation (p < 0.05) in females.

Three peptide clusters were significantly associated with age regardless of sex. Two of these clusters—clust_73 and clust_1054—originated from *collagen alpha-1(I)* and *collagen alpha-6(IV)* chains, respectively, and both demonstrated a significant negative correlation with age (Figure 7C) (Table S4). This observation aligns with previous findings by Petra Zürbig et al. [21], who also reported a decline in collagen fragment abundance with advancing age. These reductions likely reflect age-associated remodeling of the extracellular matrix, which contributes to tissue stiffening and organ dysfunction in older individuals.

**Figure 7.**
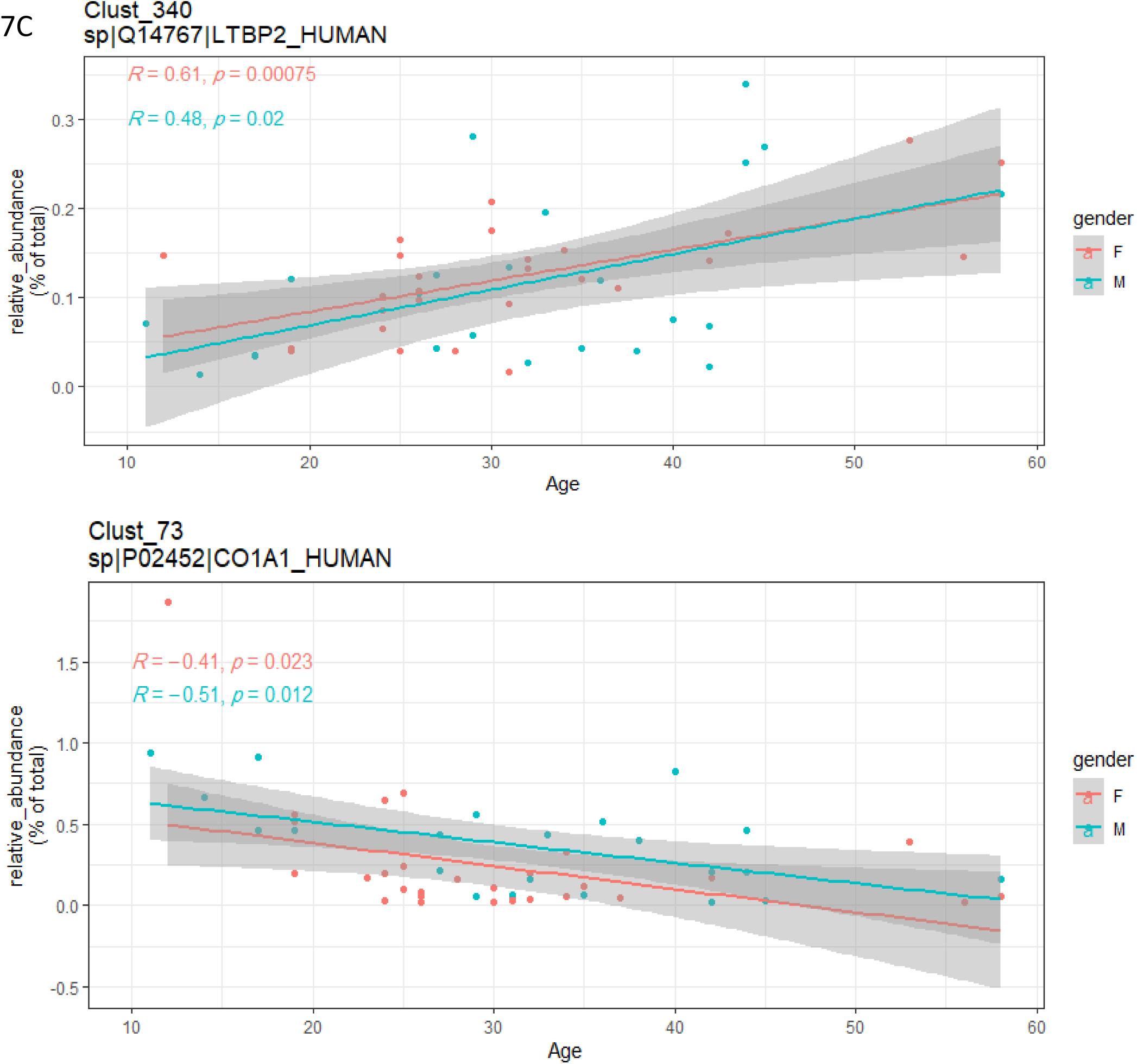
**C:** Correlation plot showing examples of age-correlated urinary peptide clusters with a significant positive or negative correlation (p < 0.05) in both males and females.

The third sex-independent cluster, clust_340, derived from *latent-transforming growth factor beta-binding protein 2*, was positively correlated with age (Figure 7C). This protein is involved in the regulation of TGF-β signaling, and its age-associated increase may reflect compensatory or pathological mechanisms related to tissue remodeling or inflammation.

We further evaluated the ageing association among the most abundant peptide clusters (relative abundance > 1%). Of these, 9 clusters showed a positive correlation with age, while only one showed a negative correlation. These high-abundance, ageing-associated peptides originated from several biologically relevant proteins, including:uromodulin, fibrinogen alpha chain, beta-defensin 1, progranulin, metallothionein-1G, albumin, insulin-like growth factor II, collagen alpha-1(XVIII) chain, and alpha-2-HS-glycoprotein.The involvement of such proteins—many of which play roles in immune regulation, extracellular matrix composition, and growth factor signaling—highlights systemic biological changes that occur with age and are reflected in the urinary peptidome.

## Materials and Methods

### Urine Sample Collection and Native Peptide Purification

Voided urine samples were obtained from 55 healthy adult volunteers, following the standardized urine collection guidelines recommended by the HUPO Human Kidney and Urine Proteome Project (HKUPP)[22]. All adult volunteers underwent regular health check-ups at Shinrakuen Hospital (Niigata Prefecture, Japan) and were confirmed to have no medical conditions requiring treatment. Protein precipitation was performed using 1 ml of frozen urine samples using the methanol/chloroform method as previously described [23]. The resulting supernatants, enriched in small endogenous peptides, were further processed as previously described[24]. All peptide extracts were aliquoted and stored at – 80 °C until further use to preserve sample integrity and prevent peptide degradation.

### LC-MS/MS Analysis

For each sample, 1 μg of purified urinary peptides was analyzed in technical duplicates using an ultrahigh-pressure nanoflow chromatography system (nanoElute, Bruker Daltonics) coupled to a trapped ion mobility quadrupole time-of-flight mass spectrometer (timsTOF Pro, Bruker Daltonics) operating in parallel accumulation serial fragmentation (PASEF) mode. Peptides were separated on a 25 cm × 75 μm inner diameter C18 column (1.6 μm particle size, Aurora Series, Ion Opticks, Australia) at a flow rate of 400 nL/min. A 70-minute linear gradient increased acetonitrile concentration from 5% to 37% (mobile phase A: water with 0.1% formic acid; mobile phase B: acetonitrile with 0.1% formic acid), and the column was maintained at 50 °C.

Mass spectra were acquired in positive ion mode, scanning from m/z 100 to 1700, with ion mobility separation from 0.6 to 1.6 Vs/cm² over a ramp time of 100 ms. Data-dependent acquisition (DDA) employed 10 PASEF MS/MS cycles per 1.1 s total cycle time, with a target intensity of 20,000 counts. Precursors reaching 20,000 intensity units were subjected to active exclusion for 0.4 min. Collision energies were dynamically adjusted from 20 to 59 eV based on ion mobility values to optimize fragmentation.

### Peptide Identification and Quantification

Raw spectral data were searched using the MASCOT search engine (v2.3.01, Matrix Science) against the Swiss-Prot human protein database. To estimate false discovery rates (FDR), spectra were also searched against a decoy database. Search parameters included no enzyme specificity, fixed carbamidomethylation of cysteine, no variable modifications, 50 ppm precursor mass tolerance, and 0.05 Da fragment mass tolerance. Identified peptides were exported at a 5% peptide-spectrum match (PSM) FDR.

### Peptide Cluster Construction

All distinct peptides identified across the 55 urine samples were assembled into peptide clusters. Clustering was based on sequence overlap, grouping overlapping peptides and their truncated variants into unified sets referred to as “peptide clusters”. Each cluster contained at least one “precursor peptide,” which could undergo N- or C-terminal truncation to yield smaller “truncated peptide” (Figure 1). This assembly approach enabled systematic analysis of peptide truncation patterns and abundance profiles.

### Peptide Cluster Quantification

Peptide cluster abundance was quantified per sample using a relative metric. Specifically, the sum of PSMs assigned to peptides within a given cluster was divided by the total PSM count for all endogenous peptides in that sample (Figure 1). For downstream analyses, only peptide clusters detected consistently in at least 50% of samples were retained, ensuring robust and systematic representation.

### Assessment of Precursor Peptide Stability

To evaluate the degradability of precursor peptides, we calculated a stability index for each cluster. The SI was derived by dividing the PSM count of precursor peptides by the total PSM count of that peptide cluster. Higher SI values indicated greater stability and lower susceptibility to proteolytic cleavage. In addition, predicted half-life and formation entropy for each precursor peptide were computed using the HLP server, providing complementary metrics of peptide stability and intrinsic disorder.

### Investigation of the differentiated peptide clusters between male and female

To identify peptide clusters differentially abundant between male and female donors, we employed the Wilcoxon rank-sum test. Fold change (FC) was computed by dividing the mean relative abundance in females by the corresponding mean in males. Only peptide clusters meeting both significance (P < 0.05) and magnitude (FC ≥ 1.5) criteria were considered sex-differentiated.

### Age Correlation Analysis

Spearman correlation analysis was performed to assess the relationship between peptide cluster abundance and donor age. Clusters with P-values < 0.05 were classified as significantly age-associated.

### Data Processing and Statistical Analysis

All data preprocessing and statistical analyses were conducted using R. The following R packages were employed: *dplyr*, *tidyr*, *stringr*, and *stringi* for data manipulation; *rstatix* for statistical tests; *ggpubr* for correlation plotting; and *ggplot2* for data visualization. In-house R scripts were developed to automate peptide assembly, peptide cluster creation, and quantification workflows.

### Biological Implications and Novel Insights

Importantly, our comprehensive analysis suggests that the urinary peptidome reflects not only general proteolytic processes but also subtle physiological differences between sexes. The finding that certain peptide clusters, such as hepcidin-25 and granulins, follow sex-dependent abundance patterns coherent with circulating protein data underlines the potential of urinary peptides as non-invasive biomarkers for systemic biological states. Furthermore, the enrichment of male-upregulated clusters in collagen-related processes raises intriguing possibilities regarding sex-specific extracellular matrix turnover, which warrants further investigation.

Additionally, the robust clustering approach applied here—assembling over 30,000 peptides into 13,163 structured clusters—provides a refined framework for peptidome interpretation. By distinguishing precursor peptides and their truncated peptide forms, we achieved a deeper understanding of peptide stability and fragmentation behavior. Our stability index analysis not only demonstrated meaningful correlations with predicted peptide half-life and entropy but also suggested that peptide intrinsic stability may contribute to their detectability in the urine matrix. This insight opens new avenues for exploring peptide resilience against endogenous peptidases, which is a crucial factor in their potential use as biomarkers or therapeutic agents.

## Conclusion

In summary, this study represents the most extensive characterization of the human urinary peptidome to date, identifying over 30,000 endogenous peptides and defining over 13,000 structured peptide clusters. Through systematic analysis, we uncovered robust sex-specific peptidomic signatures that mirror known biological differences, thereby highlighting the physiological relevance of urinary peptides. Moreover, by introducing a peptide stability index and correlating it with structural and abundance parameters, we established a novel framework to assess peptide resilience in biofluids. These findings not only enrich our understanding of urinary peptidome biology but also lay a solid foundation for future biomarker discovery and functional peptide research.

## Data Availability

All data produced in the present study are available upon reasonable request to the authors

## Institutional Review Board Statement

This study was conducted in accordance with the Declaration of Helsinki and was approved by the ethics committees of Shinrakuen Hospital (No. H290005) and Niigata University (No. 2021-0025).

## Informed Consent Statement

Urine samples used in this study were collected from left-over specimens of healthy subjects after laboratory tests for their health check at Shinrakuen Hospital and were anonymized with link information unavailable to the investigators. The ethics committees approved the collection and use of these samples by obtaining informed consent in the form of opt-out.

## Acknowledgments

COI: BBC is the Collaborative Research Laboratory of Tosoh Corporation.

---

**Table S1:**
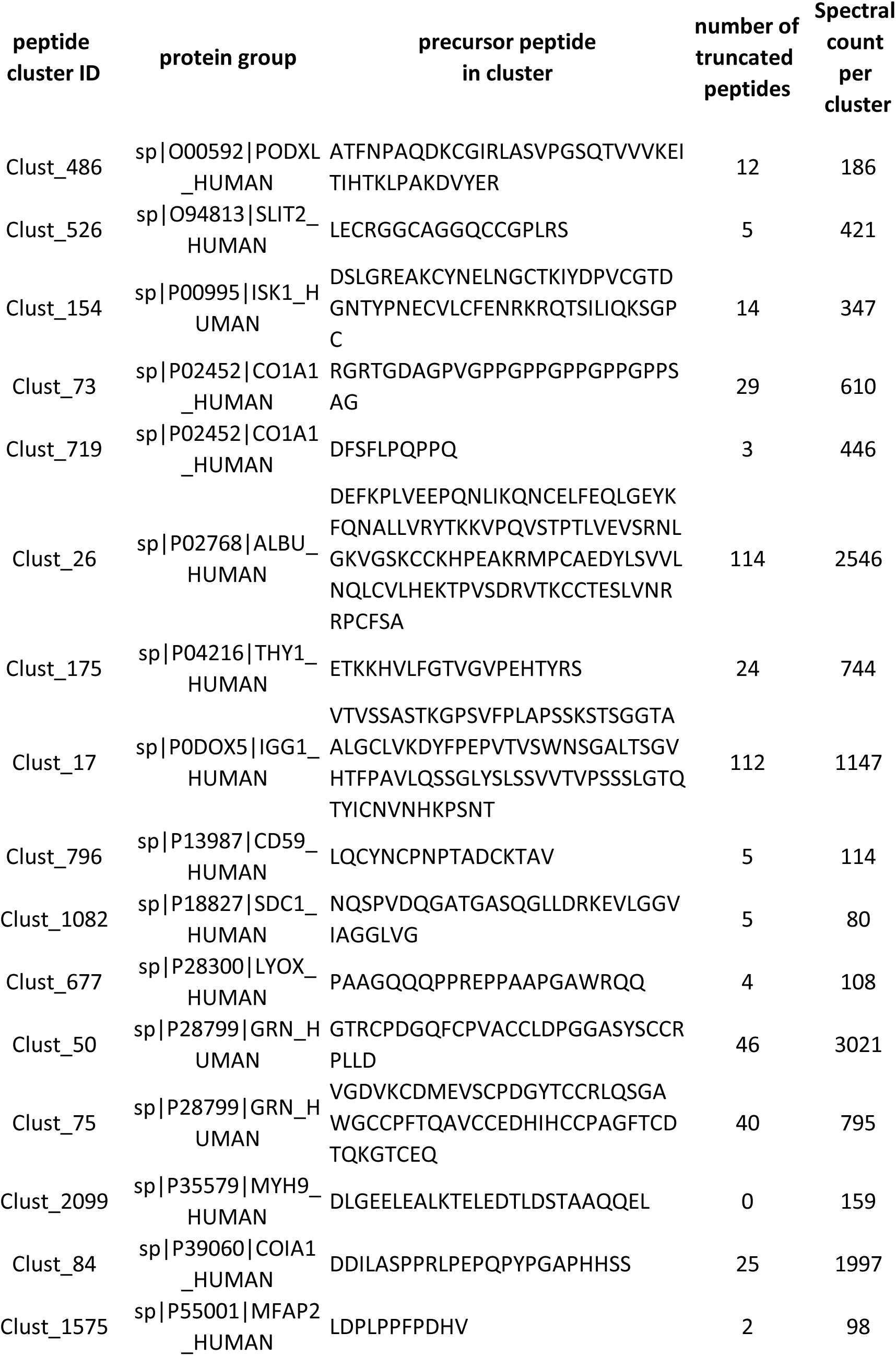
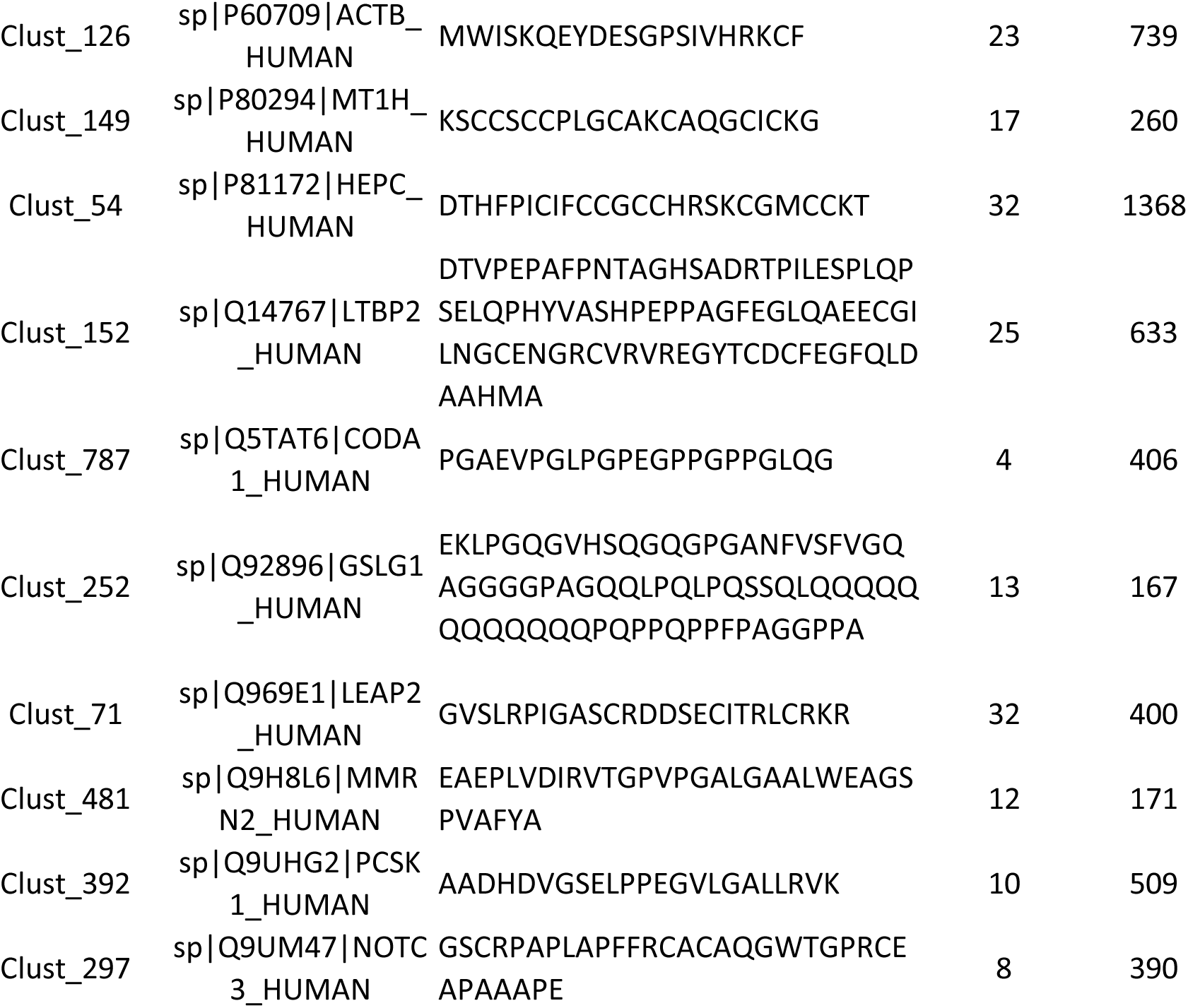
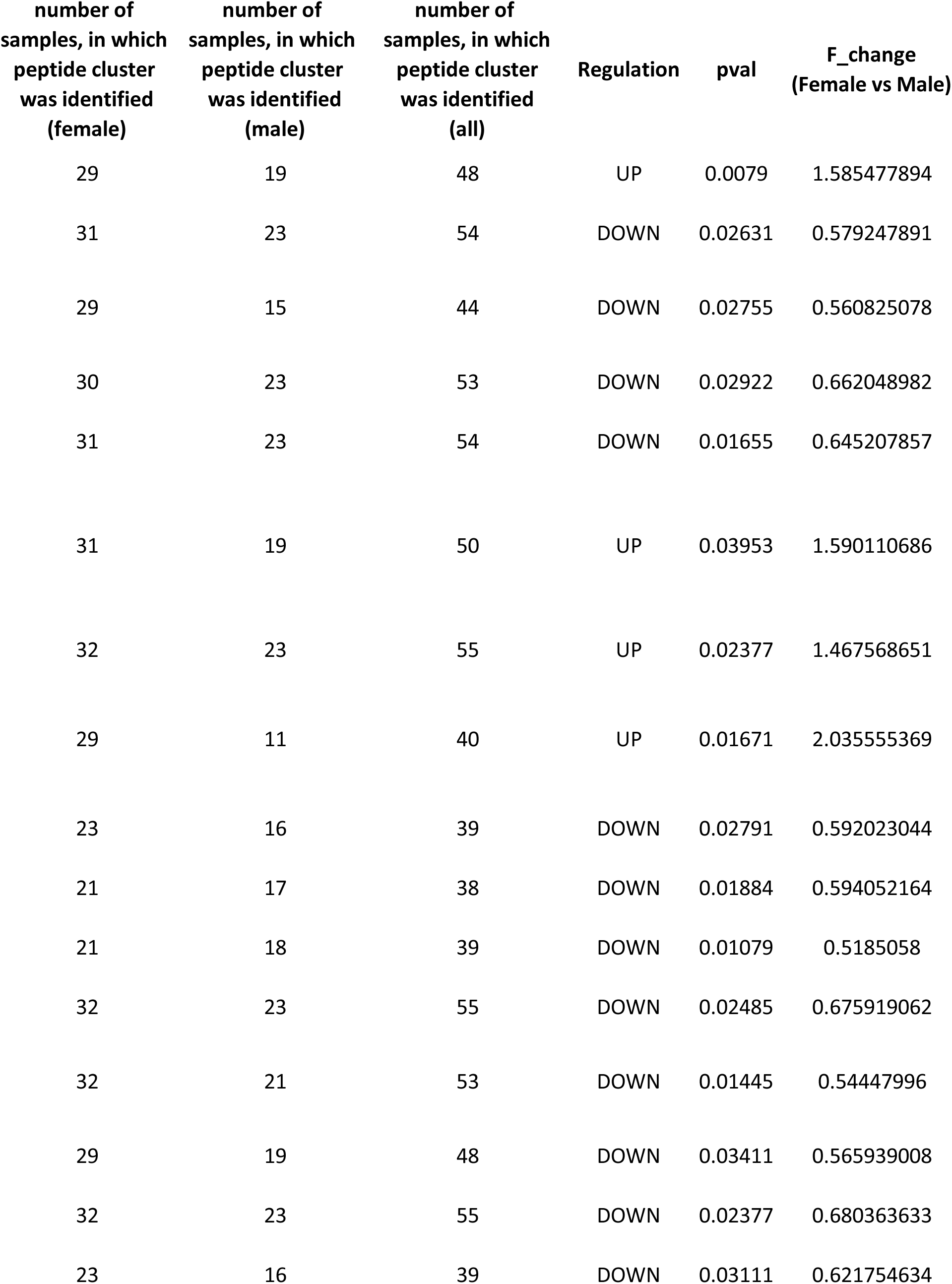
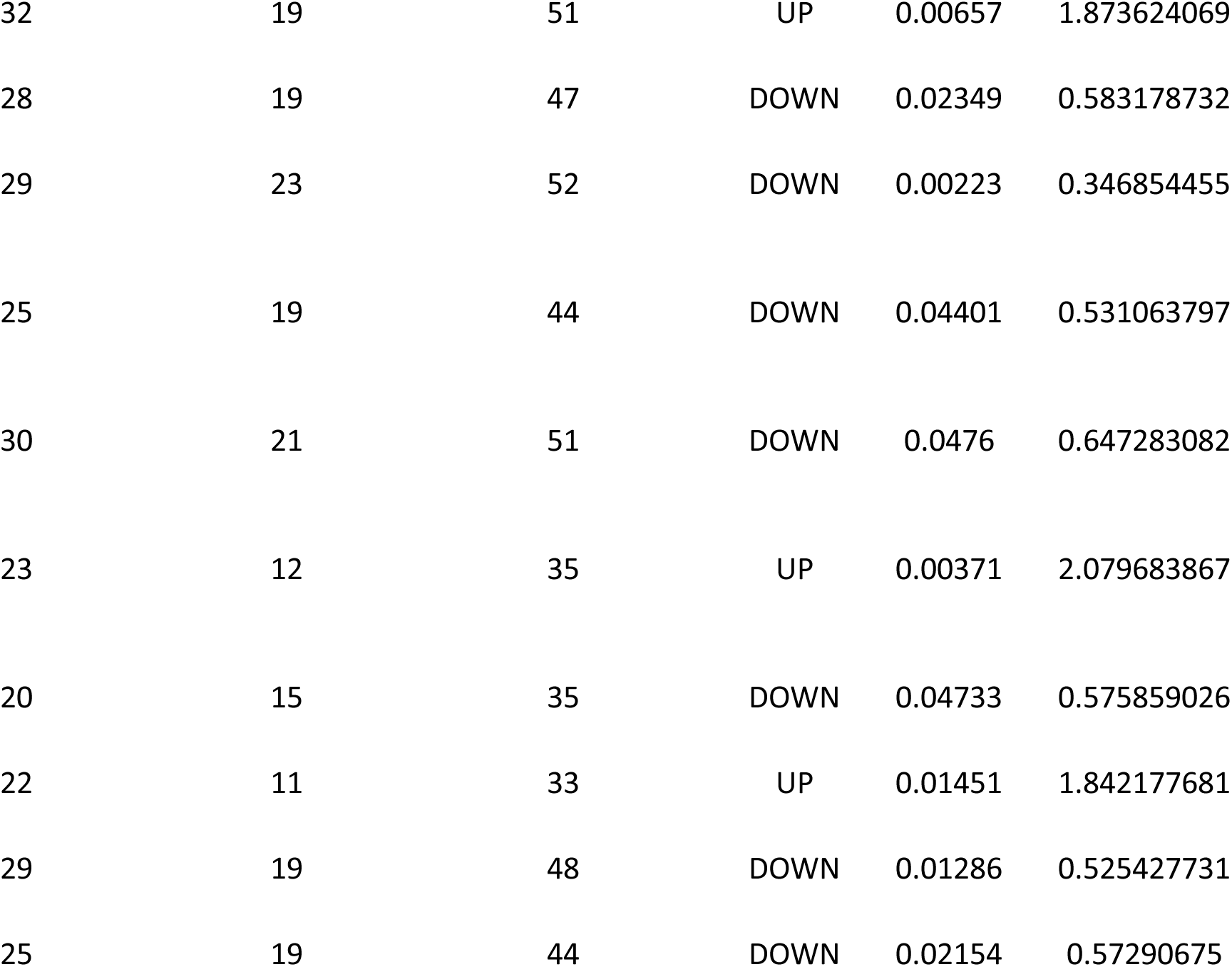
shows 26 differntiated peptide clusters between male and female (p_value < 0.05, fold change = 1.5 fold)

**Table S2:**
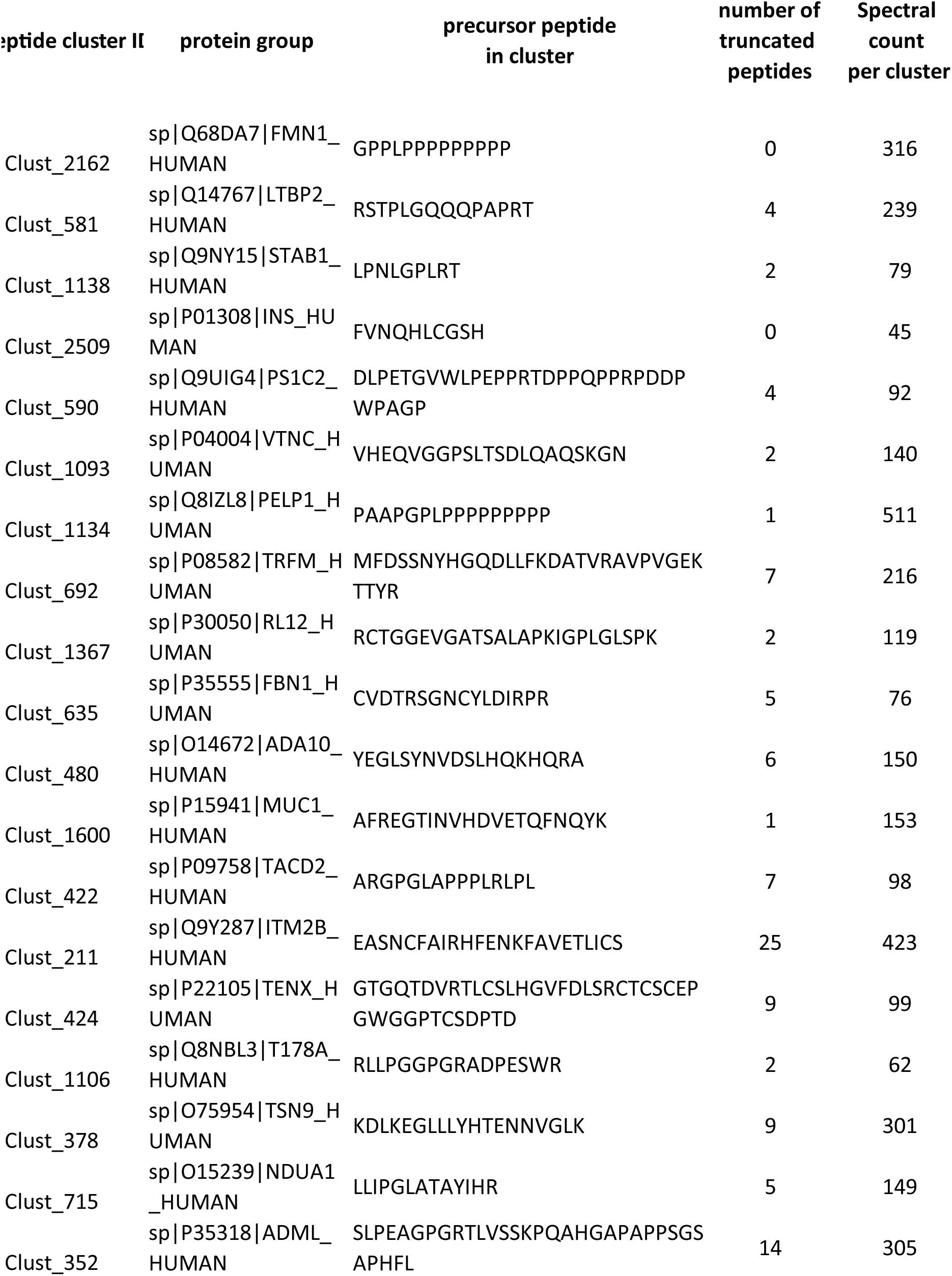
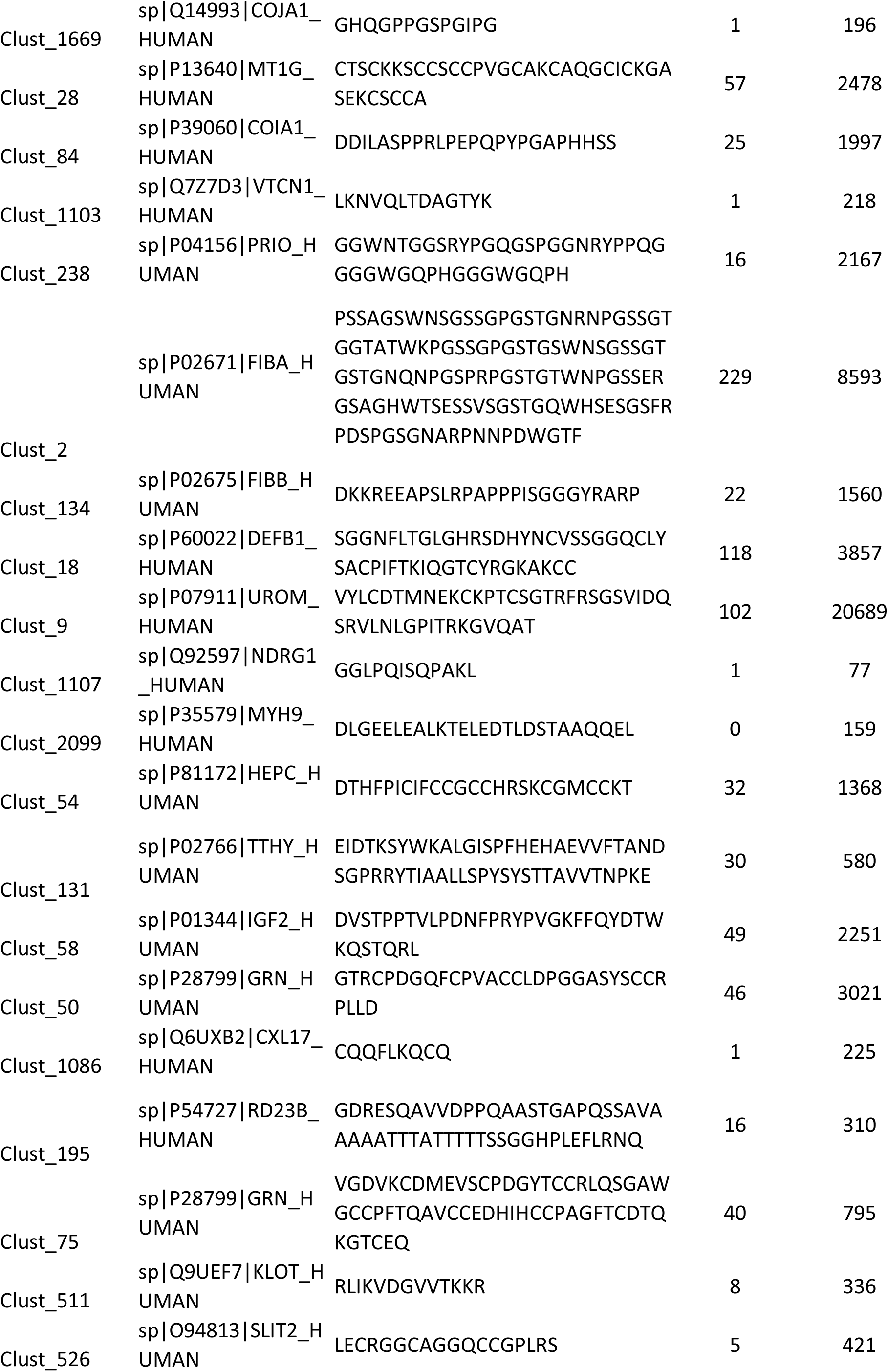
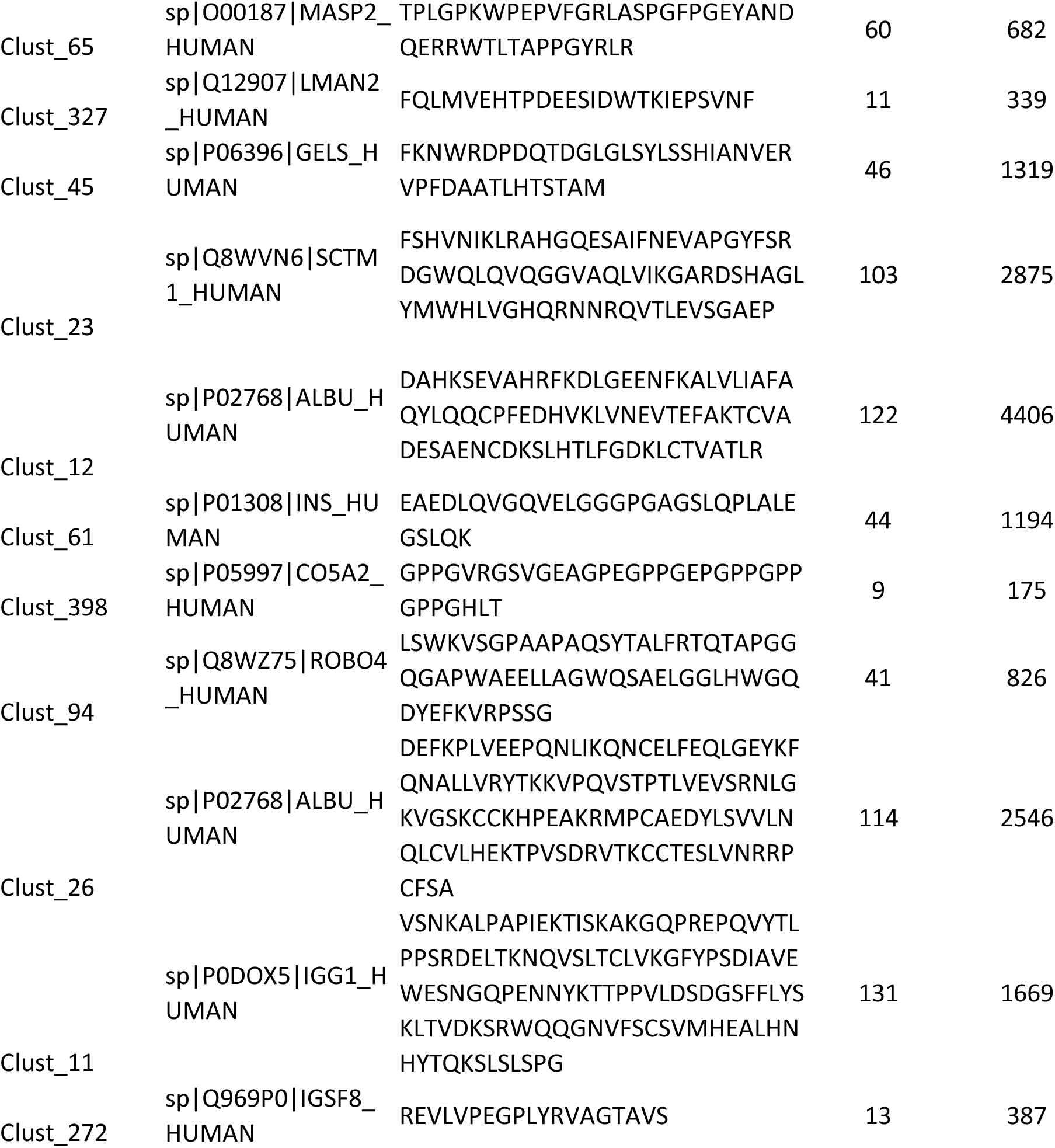
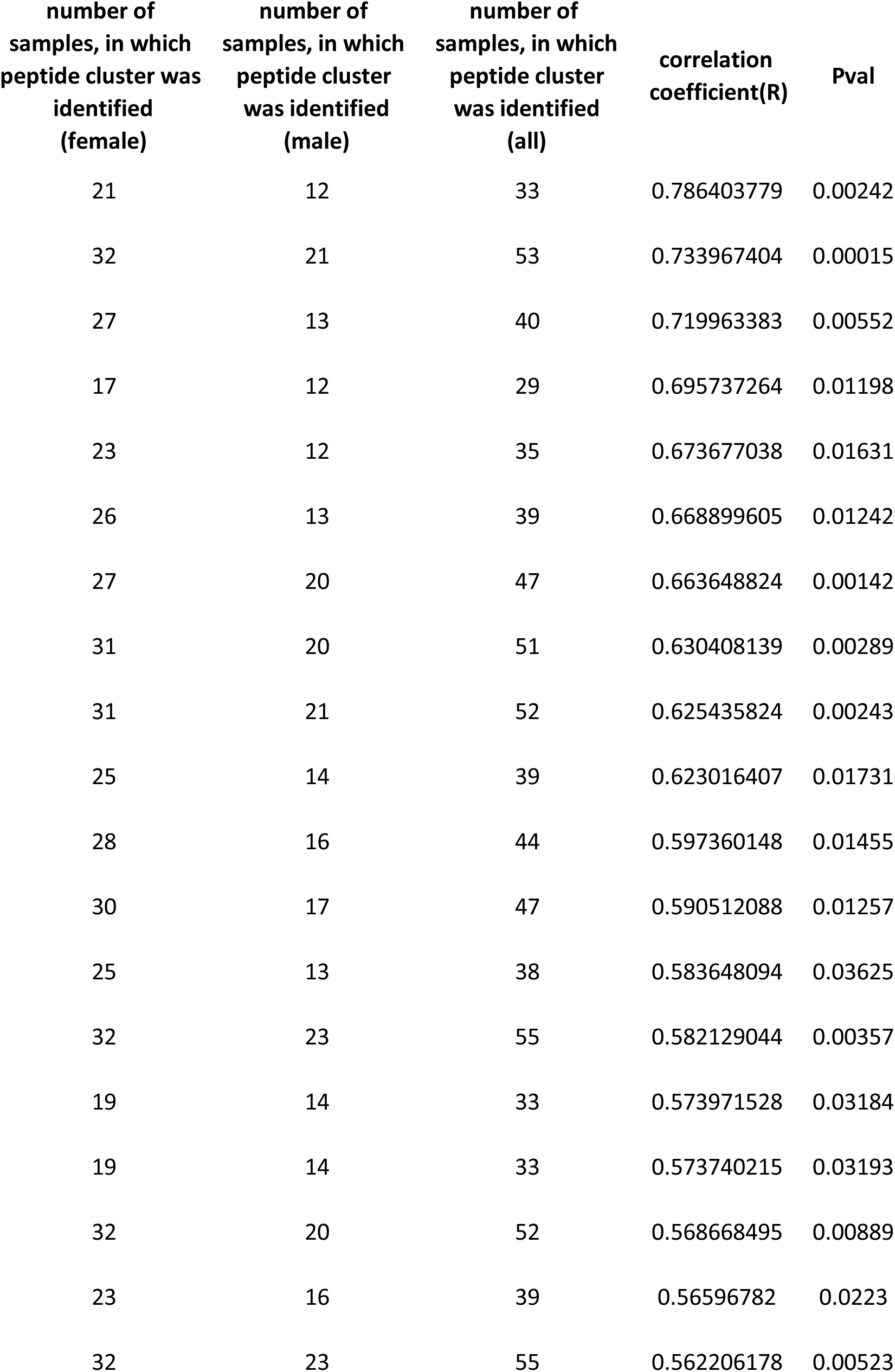
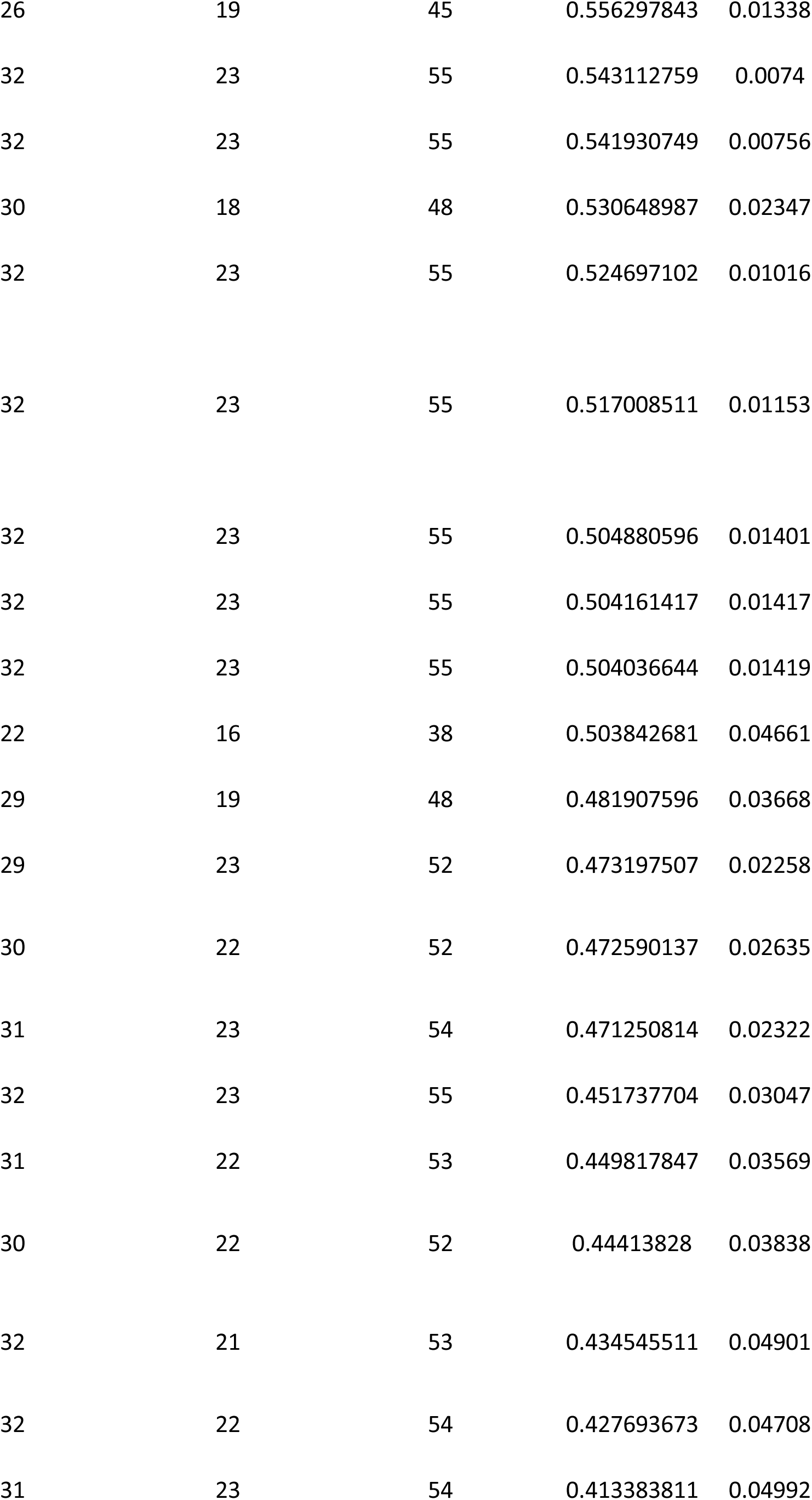
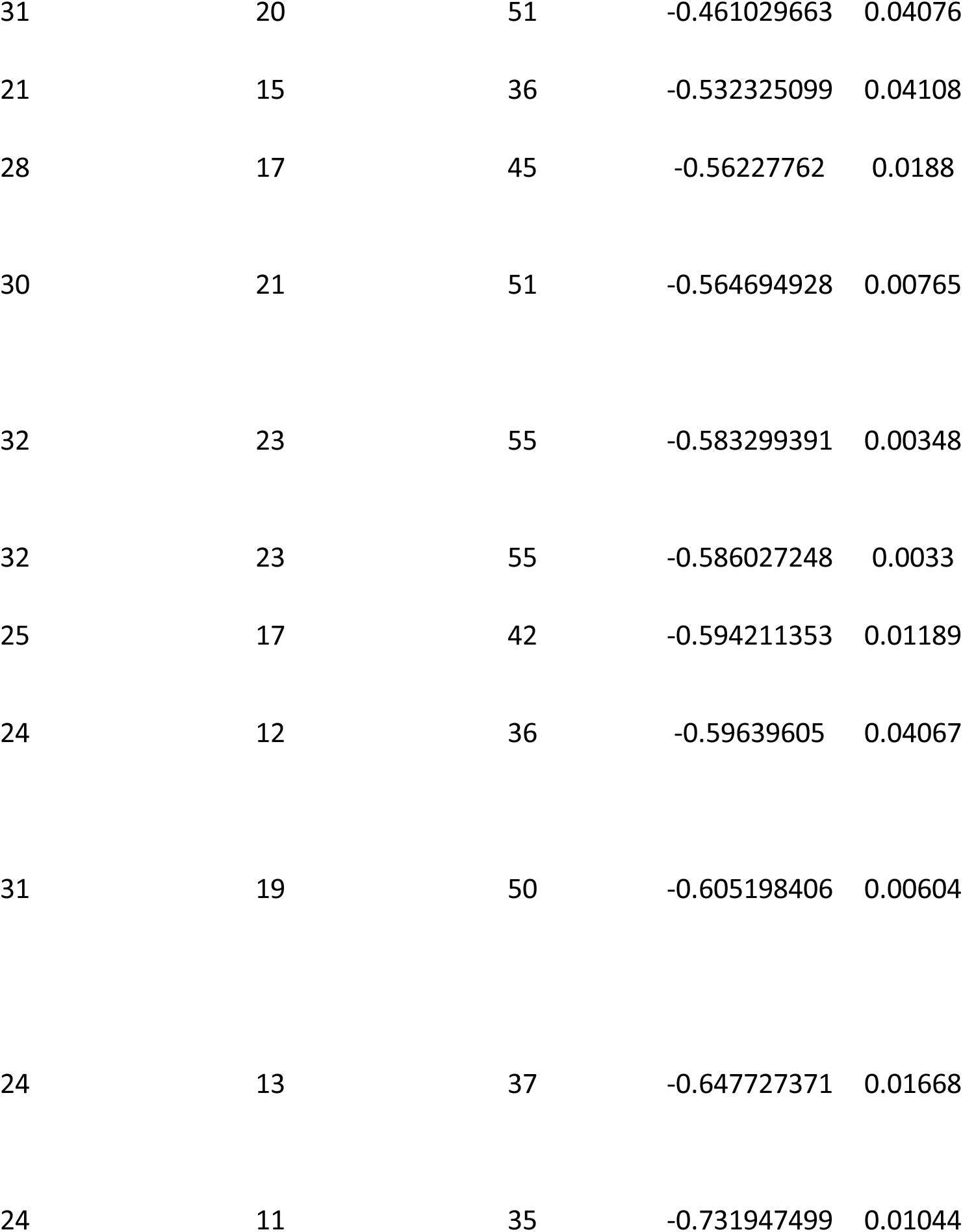
shows 50 peptide clusters that signifcantly correlated with age in male (p_value < 0.05)

**Table S3:**
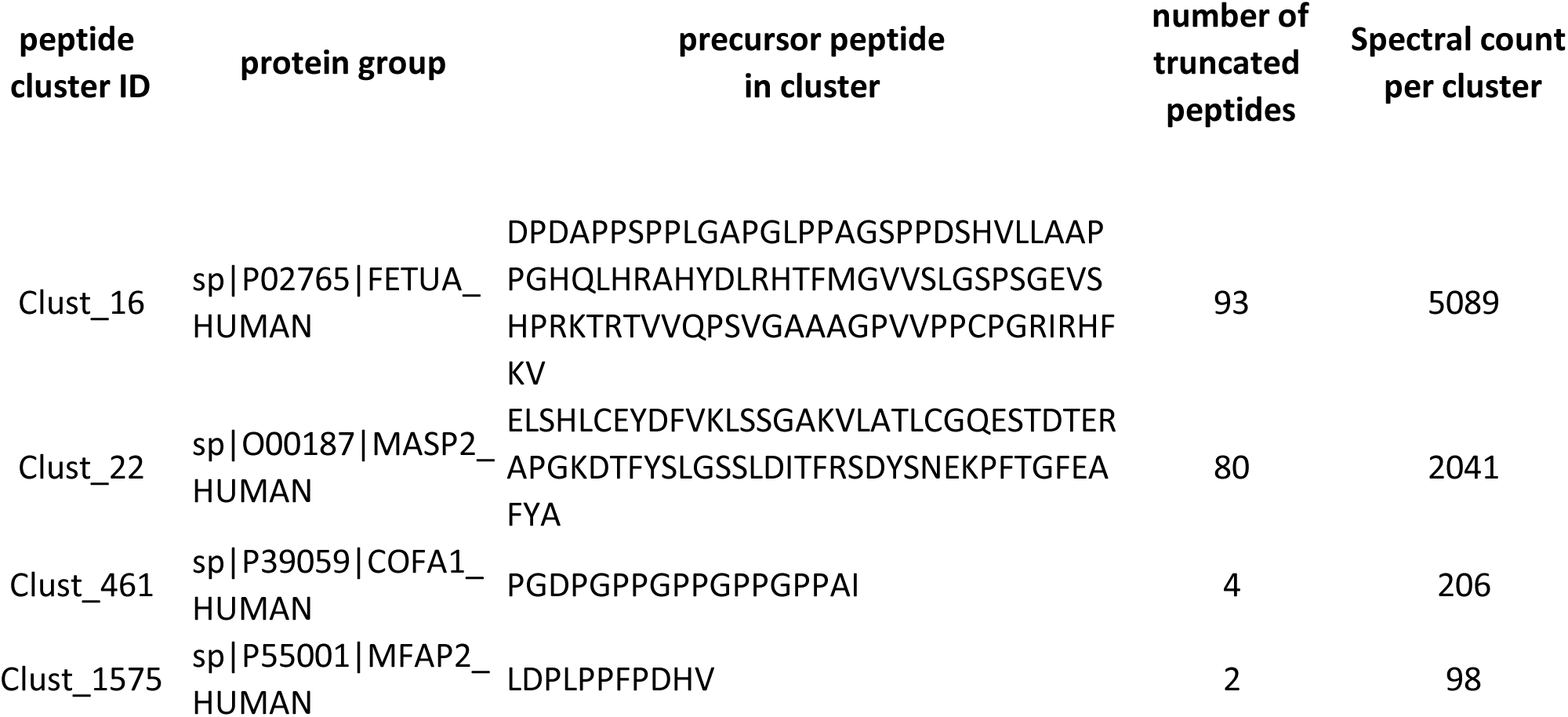
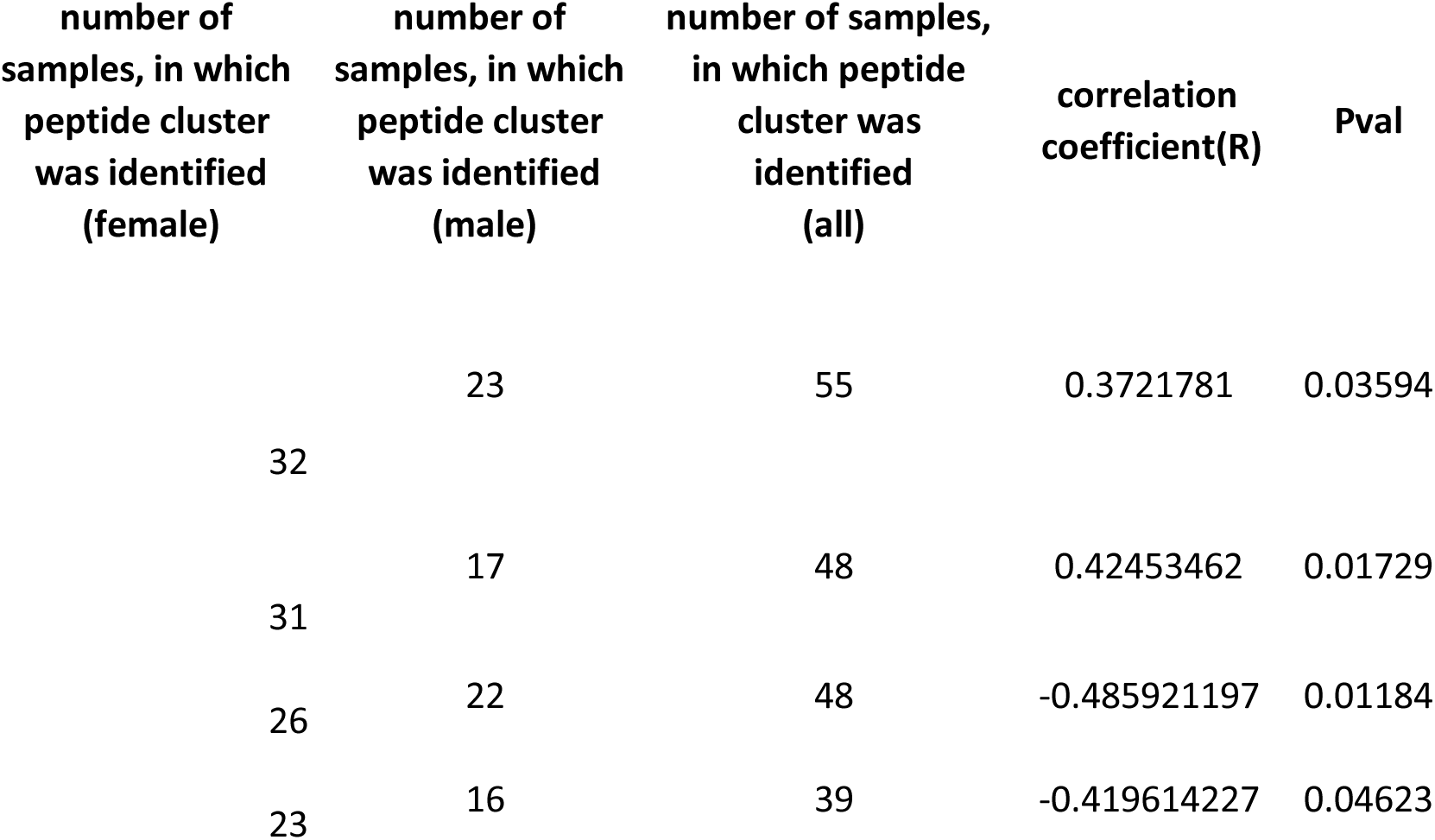
shows 4 peptide clusters that signifcantly correlated with age in female (p_value < 0.05)

**Table S4:**
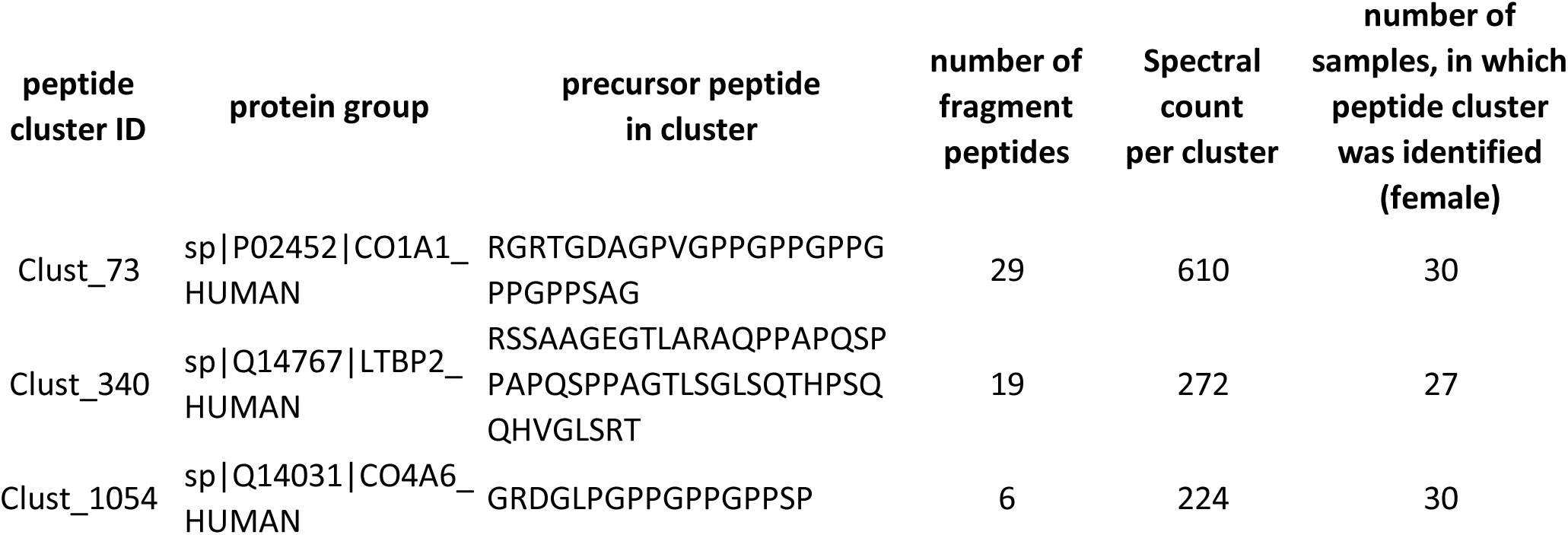
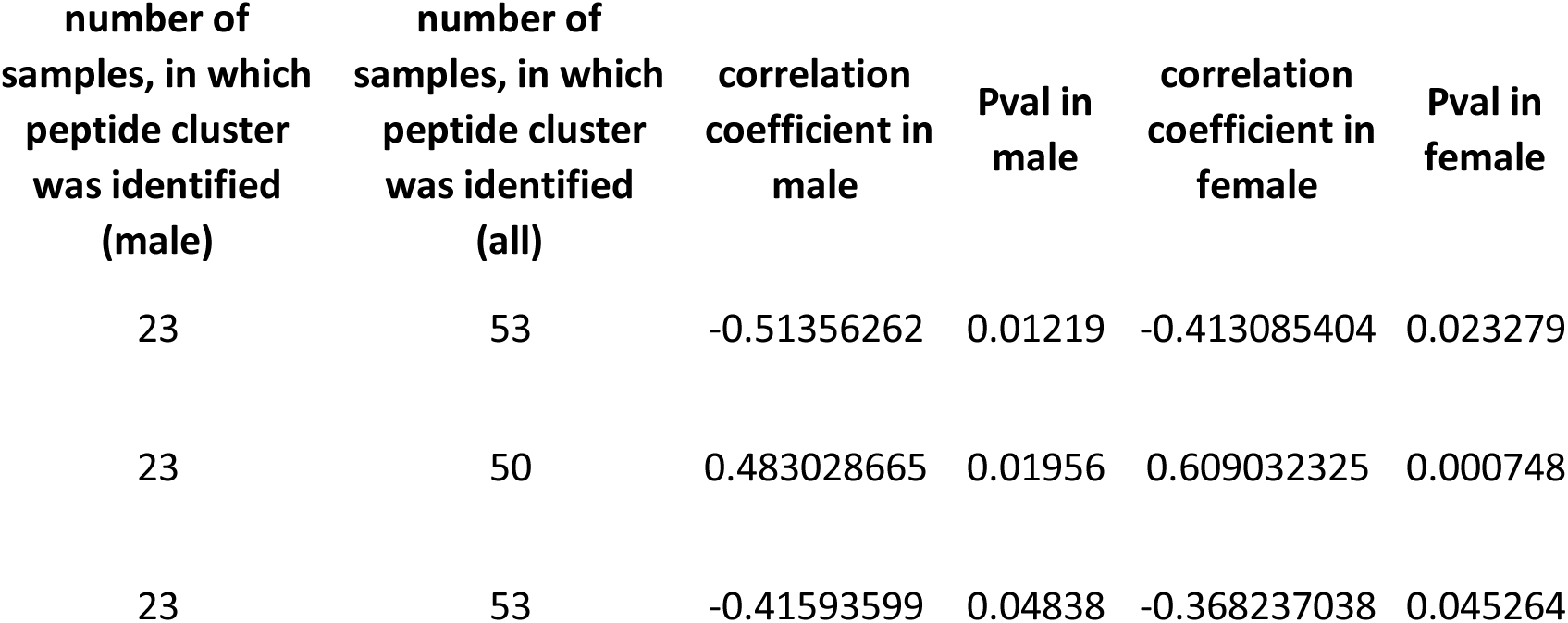
shows 3 peptide clusters that signifcantly correlated with age in male and female(p_value < 0.05)

